# Effect and acceptability of co-created interventions linking public rehabilitation programs with civil society involvement for physical activity engagement – a convergent mixed methods pilot study

**DOI:** 10.1101/2024.08.08.24311541

**Authors:** Ida Kær Thorsen, Julie Midtgaard, Marie Lønberg Hansen, Katja Thomsen, Henrik Søborg, Helle Oldrup Jensen, Thomas Peter Almdal, Janne Kunchel Lorenzen, Anders Blædel Gottlieb Hansen, Mathias Ried-Larsen

## Abstract

**Background:** Public physical activity programs are time-limited and often lacking sufficient support for citizens to maintain physical activity engagement. In this project, municipal stakeholders; civil society organizations; citizens with type 2 diabetes (T2D), cardiovascular diseases (CVD), and/or obesity; and researchers were involved in the co-creation and implementation of interventions to support citizens in continuing physical activity engagement following a municipal rehabilitation program. The primary aim of this study was to investigate the effect of these interventions on physical activity engagement in civil society sports organizations. Secondary aims were to investigate acceptability and effect of these interventions on physical and mental health, and organizational development and collaboration.

**Methods:** This was a convergent mixed methods study using a quantitative prospective intervention study with a historic control group, and a qualitative descriptive study. These were analyzed separately and then integrated. Quantitative data from citizens were collected before; and 0, 3 and 6 months after ending their rehabilitation program. Outcomes included physical activity engagement in civil society organizations; and changes in objectively measured physical activity, physical and mental health. Qualitative data were collected among citizens, civil society, and municipal representatives. Themes included physical activity engagement, acceptability, and organizational development and collaboration.

**Results:** Among the 33 included citizens (58% women, median (25^th^; 75^th^ percentile) age of 67.6 (63.9; 74.1) years), six lived with T2D; nine with CVD; and 18 were obese. Of the 21 citizens who were not engaged in physical activities in civil society organizations before entering the rehabilitation program, 67% started and remained active at 6-month follow-up—significantly more than in the historic control group. Light physical activity increased by a mean (95% confidence interval) of 15.4 (2.3; 28.5) min/day from 0 to 6 months after the rehabilitation program; all other outcomes remained unchanged. This was assisted by experienced high acceptability of the linking interventions and strengthened collaboration between the municipal health center and civil society organizations.

**Conclusions:** The co-created interventions led to increased physical activity engagement in civil society organizations. This supports co-creation as a method to link municipal rehabilitation programs with physical activities in civil society organizations.

**Trial registration:** ClinicalTrials.gov: NCT05493345, 2022-08-05

## Background

Increased physical activity levels improves the prognosis of a wide range of diseases, among those with type 2 diabetes (T2D), cardiovascular diseases (CVD), and obesity (1). In Denmark, the municipalities are obliged to offer health-related rehabilitation—however, the prescription of physical activity programs is highly variable; the programs are time-limited; and the support for continued adherence is often advice-based (2). Although people with T2D, CVD, and obesity are often motivated to continue their new active habits when their rehabilitation program ends, the provided support is insufficient in securing physical activity adherence and integration into their everyday lives (2–4). The absence of social community and perception of inaccessibility of physical activities makes engagement seem infeasible for these persons (4). Further, the transition out of rehabilitation into sustainable socially supporting structures for physical activity engagement remains unsettled (2). A potential solution is to involve civil society sports organizations in this transition (2, 5). In many Danish municipalities, civil society organizations constitute an unexploited source of social support characterized by social solidarity (6), but the collaboration between them and the public health care institutions can be complicated (7).

*Social prescribing*—as defined by the National Health Service England—involves referral from local health services to a link worker, who connects citizens with community-based activities, groups, and services (8, 9). The evidence for social prescribing is still rather sparse and varied (9). A recent review indicates that regular weekly activities and social relationships are key aspects of continued physical activity engagement in the attempt to link public rehabilitation programs with activities in civil society organizations (5). Regular weekly activities promote integration into people’s everyday lives, while social networks and easy accessibility is fostered in the transition from rehabilitation to civil society organizations (5). As such, these aspects are influenced by the collaboration between public health care institutions and civil society organizations (5, 10).

However, adapting or developing attractive activities for previously inactive persons, and identifying civil society organizations that wish to integrate new target groups may be challenging (5, 11).

In project Active Communities, we aspired to link a municipal health center with civil society organizations to increase physical activity adherence among the citizens based on a partnership between three research institutions and a Danish rural municipality (7). In the first phase of the project, we involved municipal stakeholders; civil society organizations; citizens with T2D, CVD, and/or obesity; and researchers in the co-creation of linking interventions to support citizens in continuing physical activity engagement when their rehabilitation program ends (7). In the second phase, we evaluated these linking interventions using quantitative methods to investigate the acceptability and effects of the interventions—primarily on a citizen level; qualitative methods to investigate the attitudes and perspectives towards the interventions and collaboration; and integration to provide new enhanced insights. The findings from the second phase are reported in the present paper.

The primary aim of this study is to investigate the effect of co-created linking interventions on physical activity engagement in civil society organizations. Secondary aims include investigations of 1) the acceptability of these linking interventions; 2) whether these interventions lead to maintained physical and mental health; and 3) the collaboration between and the organizational development in the municipal health center and civil society organizations.

## Methods

### Research design

This is a pilot study that constitutes the second part of Project Active Communities and was designed to evaluate the acceptability and effect of four interventions developed in the first part of the project. A program theory was developed and used as a guiding tool for the project design—this part of the project focuses on the outcomes (Figure 1). An overview of the study design, sampling sites, data collection, and analysis is presented in Figure 2. This study uses triangulation of methods, sources, and researchers in a convergent (QUAN-QUAL) mixed methods design (12). The quantitative component was a prospective intervention study with a historic control group; the qualitative component was descriptive.

**Figure 1:**
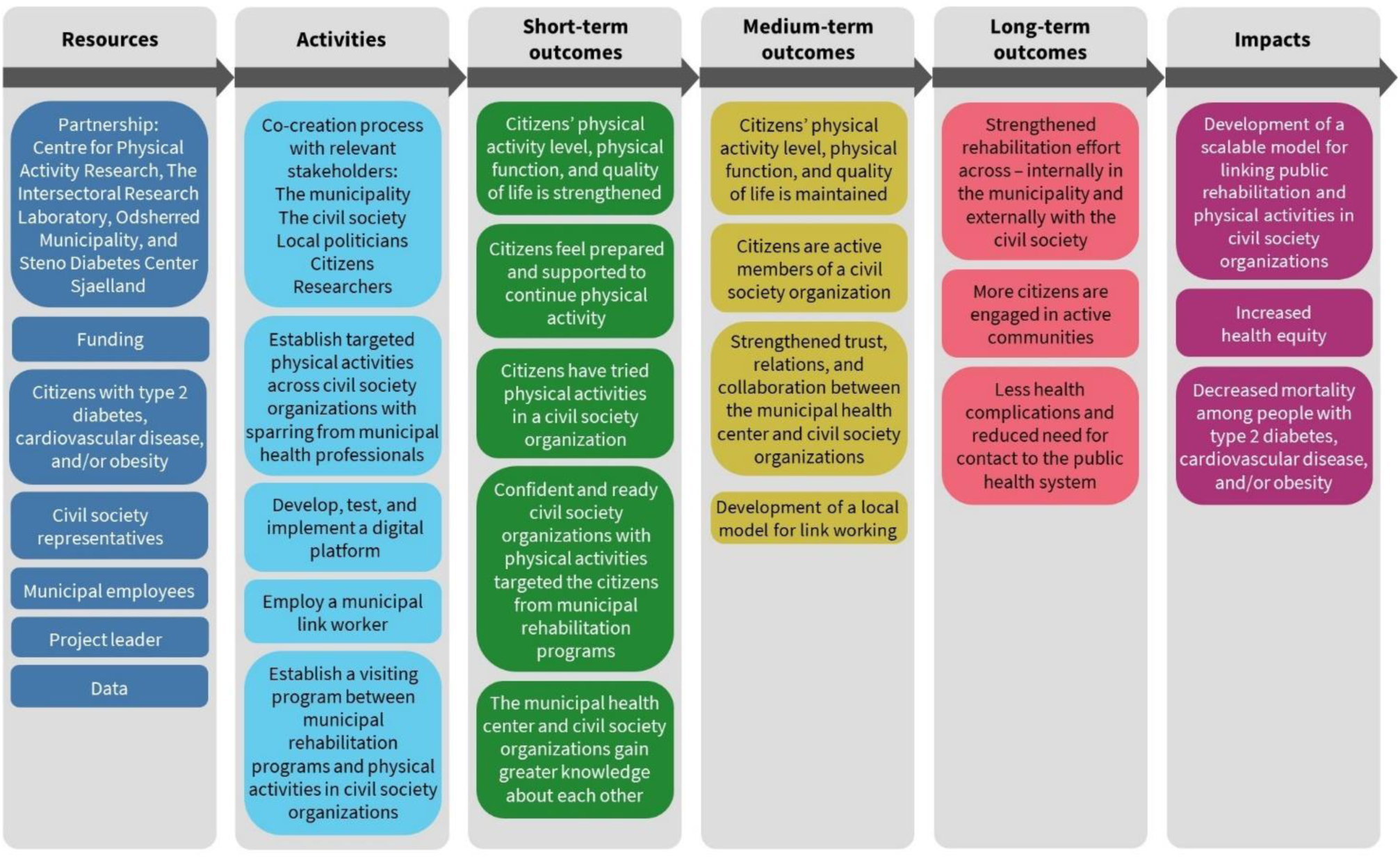
The project’s program theory, presenting resources; activities; outcomes on short-term, medium-term, and long-term; and impacts.

**Figure 2:**
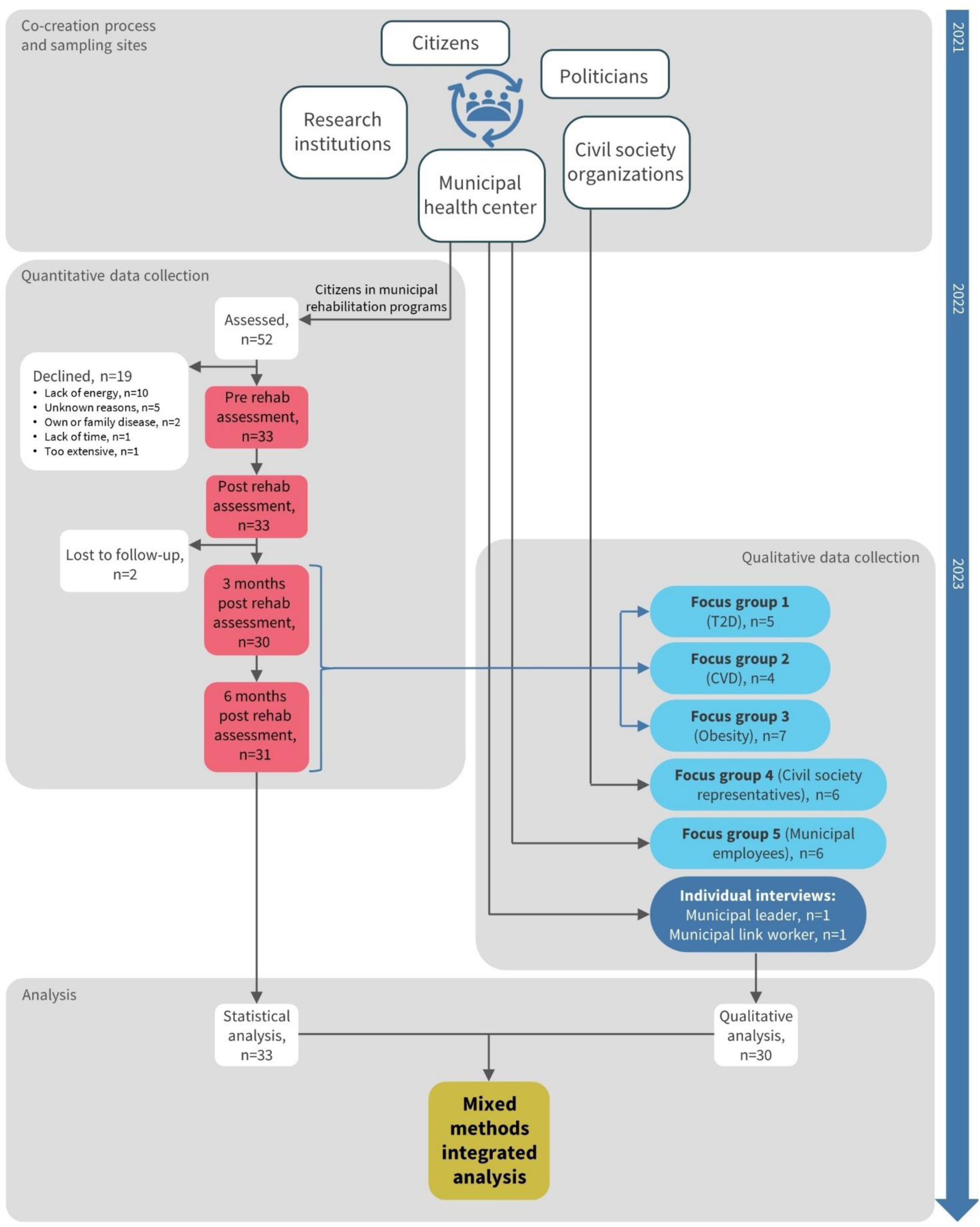
Overview of the overall study design, including sampling sites and data collection (incl. participant flow) in the quantitative and qualitative study, and the quantitative, qualitative, and integrated analysis.

The researchers’ diverse backgrounds (e.g., public health science, sports science, psychology), methods experience, and individual subjectivities were used as resources in a reflexive qualitative and integrated analysis approach. Moreover, the researcher (MLH) conducting the focus groups and individual interviews had previous interactions and collaboration with most participants from the municipality and the civil society organizations. In the first part of Project Active Communities, MLH facilitated the processes of identifying and recruiting local stakeholders; and co-creating and prototyping the linking interventions. This contributed towards a shared understanding and confidentiality between MLH and the participants in the qualitative data collection.

The intervention setting in Odsherred municipality including the health center has been described previously (7). Briefly, Odsherred is a rural municipality which is relatively poor and sparsely populated (13). In Odsherred municipality, 60.4% of the citizens are overweight (body mass index (BMI) >25) of which 25.6% are obese (BMI >30). The citizens are characterized by low socioeconomic status, and 62.4% are physically inactive (not meeting World Health Organization (WHO) recommendations) (14). Municipal health-related rehabilitation is structured in time-limited (6-12 weeks) programs, and includes education and/or practical experience with physical activity.

The study is reported according to the Mixed Methods Reporting in Rehabilitation & Health Sciences (MMR-RHS) (15).

### Participants

#### Quantitative study participants

Participants for the intervention group were recruited among citizens initiating a rehabilitation program in the municipal health center in Odsherred. All citizens who had an appointment during the period of August 5, 2022, to January 31, 2023, were approached by a health professional by telephone or face-to-face. Eligibility for participation was based on the inclusion and exclusion criteria. Inclusion criteria were ≥18 years of age; initiating a municipal rehabilitation program; and diagnosed with T2D or CVD, or being obese (body mass index (BMI)>30). Exclusion criteria were being reliant on a wheelchair. Participants in the historic control group included citizens who had prior experience with municipal rehabilitation programs (i.e., in 2019, before covid-19). They were recruited and structurally interviewed by telephone by a municipal employee. Moreover, municipal employees were recruited by email to provide data on their experience with the linking interventions.

#### Qualitative study participants

Participants for the focus groups were sampled among participants in the quantitative study by criterion-based purposeful sampling to select citizens representing T2D (Focus group 1), CVD (Focus group 2), and obesity (Focus group 3); and engagement in civil society organizations (active/not active). Participants for Focus group 4 were selected among representatives from civil society sports and patient organization involved in project Active Communities to represent diverse physical activities and organizations, while Focus group 5 included municipal employees involved in the co-creation process, and/or implementation of the interventions. All focus group participants were invited by email. The link worker and the leader of the municipal health center in Odsherred were invited by email to participate in individual interviews.

No compensation for participation was provided.

The scientific ethical committee of the Capital Region of Denmark confirmed that ethical approval was not required (22009161). All participants provided written and oral informed consent prior to participation and were assured anonymity and confidentiality.

### Interventions

Four interventions were developed and described in detail in a previously published paper (7). Briefly, the interventions include: (1) a visiting program between rehabilitation programs in the municipal health center and physical activities in civil society organizations; (2) co-created physical activities in the local community tailored to the citizens’ needs; (3) a digital platform presenting an overview of local activities for health professionals to use; and (4) a link worker employed by the municipality, among others to implement and manage the three first mentioned interventions and provide individual support for the citizens in need.

### Data collection and analysis

Quantitative data were collected from August 2022 to July 2023 and qualitative data were collected in May and June 2023; the quantitative and qualitative data were analyzed separately and then integrated (Figure 2).

#### Quantitative data collection and analysis

The participants in the intervention group reported on self-reported physical activity engagement in civil society organizations once per month from baseline to 6-month follow-up through online questionnaires sent to the participants’ email addresses. Based on the question, “During the last month, have you engaged in one or more physical activities in civil society organizations?”, participants were divided into 1. *Active in organization* covering citizens reporting engagement in physical activities in civil society organizations at pre-baseline; 2. *Not active in organization* covering citizens reporting no engagement in physical activities in civil society organizations at pre-baseline and 6-month follow-up; and 3. *New in organization* covering citizens reporting no engagement in physical activities in civil society organizations at pre-baseline and engagement in physical activities in civil society organizations at 6-month follow-up.

In addition, the historic control group provided retrospective data on physical activity engagement in civil society organizations before and after their municipal rehabilitation program. Based on the questions, A) “Think back to 2019—when your municipal rehabilitation program ended, did you continue being physically activity in a civil society organization?” and B) “Think back to 2019 again—were you physically active in a civil society organization before starting your municipal rehabilitation program?”, citizens were divided into 1. *Active in organization* covering citizens answering “Yes” to question B; 2. *Not active in organization* covering citizens answering “No” to questions A and B; and 3. *New in organization* covering citizens answering “Yes” to question A and “No” to question B.

Quantitative acceptability outcomes related to the intervention group and municipal employees’ familiarity and perception of the link worker function, the visiting program and co-created activities, and the digital platform were assessed using questions developed by the project group (Appendix 1).

Among participants in the intervention group, objectively measured physical activity; physical performance; body composition; and self-reported quality of life, well-being, stress, social isolation, and loneliness were assessed before entering a rehabilitation program (pre-baseline), immediately after ending the program (baseline), and 3 and 6 months after ending the program (3- and 6-month follow-up, respectively). The primary timeframe was baseline to 6-month follow-up; secondary timeframes include baseline to 3-month follow-up, and pre-baseline to baseline and 6-month follow-up, respectively.

Objectively measured physical activity outcomes include moderate-and-vigorous physical activity (MVPA) time, light physical activity (LPA) time, sitting time, total physical activity, and steps. These were assessed using accelerometers (AX3, Axivity) attached to participants’ thigh and back and worn for 7 consecutive days. OmGui (version 1.0.0.43) were used to setup and download raw data (cwa files). Raw data were processed into physical activity intensities and types using the custom-build R package PhysAccel, while steps were processed using ActiLife (version 6.11.9). MVPA time (≥1952 counts per minute (CPM)), LPA time (≥100 and <1952 CPM), total physical activity (TPA; CPM) and steps (n/day) were assessed by the accelerometer placed on participants’ back, while sitting time (min/day) was assessed by the accelerometer placed on participants’ thigh, as described elsewhere (16).

Six-minute walk distance was assessed using the 6-minute walk test (17) and standing balance were assessed using the Tandem balance test (18). Body composition outcomes include body weight, fat percentage, and muscle mass. These were assessed using a dual frequency body composition analyzer (Tanita DC-430MA or BC-420MA).

Self-reported outcomes were assessed using validated questionnaires: physical and mental health-related quality of life was assessed using the 12-item short-form health survey (SF-12) (19); mental well-being was assessed using the 5-item WHO well-being index (WHO-5) (20); stress was assessed using the perceived stress scale (PSS) (21); social isolation was assessed using a modified version of the Valtorta index (14, 22); and loneliness was assessed using the three-item loneliness index (T-ILS) (23).

As this is a pilot study, no formal sample size calculation was performed, and thus the sample size was determined by the intervention settings that permitted recruitment until January 31, 2023. Acceptability outcome results related to citizens’ and municipal employees’ experience and satisfaction with the linking interventions will be reported descriptively as number of total participants (n of N).

Tandem balance score was analyzed using Wilcoxon signed rank test. All other continuous outcomes were analyzed using repeated measures mixed linear models including fixed effect factors for time (four levels: pre-baseline, baseline, 3-month follow-up, and 6-month follow-up) and random effects for participant identification number. Model assumptions were investigated, including 1) linearity, 2) normality of residuals, 3) homogeneity of residuals variance, and 4) independence of residual error. Standardized mean differences with 95% confidence intervals (CI) were calculated using Cohen’s d. Results are reported as standardized mean differences and least square mean differences between time points with associated 95% CIs. Categorical outcomes (i.e., social isolation and severe loneliness) were analyzed using Fischer exact test. Data were analyzed as observed. All 95% CIs and P values are two-sided. All statistical analyses were performed using Stata/SE 18 and R 4.3.0.

#### Qualitative data collection and analysis

Data were collected through focus groups and semi-structured individual interviews. The focus groups were based on a focus group schedule that included presentation of constructed statements developed based on the project’s program theory to inspire conversation and discussion among participants. The focus group method was chosen to provide insights from multiple perspectives and attitudes. The individual interviews were based on an interview schedule also developed based on the project’s program theory to provide in-depth insights to the participants experiences and perspectives. Both focus groups and individual interviews were digitally recorded, and field notes were made during and immediately after each.

Digital recordings of interviews and focus groups were transcribed and thematically analyzed using a deductive approach. The data analysis began with overall color coding of the transcribed text into overall supporting and undermining factors regarding the link between rehabilitation programs and physical activities in the civil society. This color coding was maintained when the text was subsequently coded according to the outcomes described in the project program theory: 1) Visiting program and co-created activities; 2) Digital platform; 3) Link worker; 4) Citizens’ engagement in physical activity; 5) Citizens’ physical and mental health; 6) Collaboration between the municipality and the civil society; and 7) Organizational development. Further text condensation, discussion and review of these themes led to merging of themes 1-3 and themes 4-5 into two themes covering Acceptability of the interventions and Physical activity engagement, respectively, resulting in a total of four main themes. Information power was achieved through diverse participant characteristics and a focused research aim, ensuring a comprehensive understanding (24).

#### Mixed methods integration

The quantitative and qualitative analysis were performed in parallel and then integrated through embedding methods (12). The integrated analysis will be presented partly through a joint display and partly through narrative using a weaving approach. The quantitative outcomes, qualitative themes, and reporting approach for integration are described in sTable 1. The results are reported in six subsections: 1) Participant characteristics, using descriptive statistics; 2) Physical activity engagement, using joint display; 3) Acceptability of the interventions, using integration through narrative; 4) Citizens’ physical and mental health, using quantitative results only; and 5) Collaboration between the municipality and the civil society, and 6) Organizational development— 5 and 6 using qualitative results only.

## Results

### Participant characteristics

Between August 2022 and January 2023, 52 persons initiating a rehabilitation program were invited to participate in the intervention study (Figure 2). Of these, 19 declined to participate. Of the 33 persons enrolled in the intervention group, a total of 31 participants completed the 6-month follow-up; six of these had T2D, nine had CVD, and 18 were obese. From these groups, five of six invited participants with T2D, four of seven invited participants with CVD, and seven of nine invited participants with obesity participated in homogenous focus groups (Focus groups 1-3). Characteristics of citizen participants in the quantitative and qualitative study are presented in Table 1. In the intervention group, the median (25^th^; 75^th^ quartile) age was 67.6 (63.9; 74.1) years, and 18 (57.6%) were women and 14 (42.4%) were men. At baseline, participants spent a median (25^th^; 75^th^ quartile) of 28.1 (10.7; 49.4) min/day in MVPA. The historic control group contained relatively more men and less citizens living alone compared with the participants in the intervention group (untested). In Focus groups 4 and 5, respectively, 6 of 8 of the invited civil society representatives and 6 of 6 of the invited municipal employees participated. The civil society representatives were from patient associations representing T2D (n=1) and CVD (n=1); and civil society sports organizations, including a gymnastics association (n=1), a fitness association (n=1), and a soccer association (n=1). The municipal employees were physiotherapists (n=3), nurses (n=2), and a consultant on civil society organizations (n=1).

**Table 1:** Sociodemographic and disease-related citizen participant characteristics.

|  | Intervention group | Historic control group | Focus group 1: Citizens with T2D | Focus group 2: Citizens with CVD | Focus group 3: Citizens with Obesity |
| --- | --- | --- | --- | --- | --- |
| **N** | 33 | 51 | 5 | 4 | 7 |
| **Rehabilitation program<sup>a</sup>** |  |  |  |  |  |
| Type 2 diabetes | 6 (18.2) | 5 (9.8) | 5 (100.0) | 0 (0.0) | 0 (0.0) |
| Cardiovascular disease | 9 (27.3) | 13 (25.5) | 0 (0.0) | 4 (100.0) | 0 (0.0) |
| Overweight | 18 (54.5) | 33 (64.7) | 0 (0.0) | 0 (0.0) | 7 (100.0) |
| **Already engaged in physical activities in a civil society organization<sup>a</sup>** |  |  |  |  |  |
| Yes | 12 (36.4) | 36 (70.6) | 4 (80.0) | 1 (25.0) | 1 (14.3) |
| No | 21 (63.6) | 15 (29.4) | 1 (20.0) | 3 (75.0) | 6 (85.7) |
| **Sex<sup>a</sup>** |  |  |  |  |  |
| Women | 19 (57.6) | 17 (33.3) | 4 (80.0) | 0 (0.0) | 6 (85.7) |
| Men | 14 (42.4) | 34 (66.7) | 1 (20.0) | 4 (100.0) | 1 (14.3) |
| **Age (years)<sup>b</sup>** | 67.6 (63.9; 74.1) | 70.2 (67.3; 76.9) | 65-77 | 66-76 | 43-77 |
| **Highest attained level of education<sup>a</sup>** |  |  |  |  |  |
| ISCED-2011 levels 0-4 | 25 (75.8) | 31 (60.8) | 3 (60.0) | 3 (75.0) | 6 (85.7) |
| ISCED-2011 levels 5-8 | 8 (24.2) | 20 (39.2) | 2 (40.0) | 1 (25.0) | 1 (14.3) |
| **Source of income<sup>a</sup>** |  |  |  |  |  |
| Salary | 6 (18.2) | 5 (9.8) | 0 (0.0) | 1 (25.0) | 1 (14.3) |
| Retirement pension | 24 (72.7) | 40 (78.4) | 4 (80.0) | 3 (75.0) | 4 (57.1) |
| Public income support | 3 (9.1) | 6 (11.8) | 1 (20.0) | 0 (0.0) | 2 (28.6) |
| **Cohabitation status<sup>a</sup>** |  |  |  |  |  |
| Living alone | 17 (51.5) | 12 (23.5) | 3 (60.0) | 0 (0.0) | 4 (57.1) |
| Living with spouse and/or children | 16 (48.5) | 39 (76.5) | 2 (40.0) | 4 (100.0) | 3 (42.9) |
| **Chronic conditions<sup>a</sup>** |  |  |  |  |  |
| One | 4 (12.1) | n/a | 2 (40.0) | 0 (0.0) | 0 (0.0) |
| Two or more | 29 (87.9) | n/a | 3 (60.0) | 4 (100.0) | 7 (100.0) |
| **Smoking habits<sup>a</sup>** |  |  |  |  |  |
| Current | 2 (6.1) | n/a | 0 (0.0) | 0 (0.0) | 1 (14.2) |
| Earlier | 18 (54.5) | n/a | 3 (60.0) | 3 (75.0) | 3 (42.9) |
| Never | 13 (39.4) | n/a | 2 (40.0) | 1 (25.0) | 3 (42.9) |
| **Alcohol consumption<sup>a</sup>** |  |  |  |  |  |
| No alcohol | 11 (34.4) | n/a | 0 (0.0) | 1 (25.0) | 4 (57.1) |
| According to recommendations | 18 (56.2) | n/a | 5 (100.0) | 2 (50.0) | 1 (14.3) |
| Above recommendations | 3 (9.4) | n/a | 0 (0.0) | 1 (25.0) | 2 (28.6) |
| **Physical activity and function<sup>b</sup>** |  |  |  |  |  |
| MVPA time (minutes/day) | 28.1 (10.7; 49.4) | n/a | 59.5 (50.5; 75.1) | 13.1 (0.8; 26.4) | 47.5 (20.5; 69.7) |
| LPA time (minutes/day) | 166.5 (129.1; 207.8) | n/a | 195.7 (156.4; 245.3) | 131.4 (95.5; 185.6) | 180.9 (166.3; 214.0) |
| Sitting time (minutes/day) | 594.3 (530.1; 709.8) | n/a | 540.4 (488.0; 616.8) | 593.0 (530.1; 663.8) | 614.9 (515.5; 672.2) |
| Total physical activity (CPM) | 151.1 (83.0; 205.9) | n/a | 249.0 (219.6; 273.4) | 87.9 (34.6; 151.1) | 191.0 (125.2; 271.2) |
| Steps (n/day) | 6376 (3944; 7991) | n/a | 7214 (6407; 10297) | 3944 (2883; 5732) | 7176 (6445; 10326) |
| 6-minute walk test (meters) | 429 (362; 481) | n/a | 495 (475; 500) | 463 (440; 559) | 421 (394; 492) |
| Tandem balance test (score 0-30) | 30 (30; 30) | n/a | 30 (30; 30) | 30 (22; 30) | 30 (23; 30) |
| **Body composition<sup>b</sup>** |  |  |  |  |  |
| Body weight (kg) | 94.4 (75.7; 109.9) | n/a | 65.3 (63.1; 73.3) | 96.8 (93.1; 108.6) | 85.1 (77.6; 100.4) |
| Fat percentage (%) | 37.7 (30.5; 42.3) | n/a | 32.5 (30.5; 33.0) | 30.4 (20.7; 32.6) | 41.0 (37.5; 42.3) |
| Muscle mass (kg) | 51.2 (46.6; 61.6) | n/a | 41.7 (41.6; 43.2) | 62.0 (61.6; 81.9) | 47.6 (46.2; 50.5) |
| BMI (kg/m<sup>2</sup>) | 30.8 (28.1; 38.3) | n/a | 26.0 (21.3; 26.5) | 32.0 (28.7; 34.3) | 29.3 (28.8; 38.3) |
| **Self-reported measures** |  |  |  |  |  |
| SF-12 Physical Component Summary (score 0-100)<sup>b</sup> | 40.3 (30.0; 47.5) | n/a | 49.2 (45.1; 50.9) | 46.7 (43.1; 49.0) | 28.8 (27.3; 34.2) |
| SF-12 Mental Component Summary (score 0-100)<sup>b</sup> | 55.4 (49.3; 59.4) | n/a | 54.2 (50.1; 60.2) | 59.3 (57.2; 61.2) | 57.5 (50.2; 65.3) |
| WHO-5 Well-Being Index (score 0-100)<sup>b</sup> | 72 (54; 80) | n/a | 80 (76; 80) | 80 (68; 86) | 56 (40; 68) |
| Risk of depression (WHO-5 score <50)<sup>a</sup> | 4 (12.5) | n/a | 0 (0.0) | 0 (0.0) | 2 (28.6) |
| Perceived stress scale (score 0-40)<sup>b</sup> | 13 (10; 18) | n/a | 4 (1;11) | 7 (5; 10) | 13 (11; 16) |
| Social isolation (modified Valtorta score ≥4)<sup>a</sup> | 4 (13.8) | n/a | 0 (0.0) | 0 (0.0) | 3 (42.9) |
| Severe loneliness (T-ILS score ≥7)<sup>a</sup> | 1 (3.0) | n/a | 0 (0.0) | 0 (0.0) | 1 (14.3) |

Data are presented as <sup>a</sup>n (%) or <sup>b</sup>median (25<sup>th</sup>; 75<sup>th</sup> percentile). ISCED-2011, International Standard Classification of Education 2011; n/a, not available; MVPA, moderate-and-vigorous physical activity; LPA, light physical activity; TPA, total physical activity; CPM, counts per minute; BMI, body mass index; SF-12, 12-item short-form health survey; WHO, World Health Organization.

### Physical activity engagement

Figure 3 presents the results of the quantitative, qualitative, and integrated analyses of physical activity engagement in civil society organizations, including quantitative comparison with the historic control group. The experience of the 12 citizens who were already active in civil society organizations was characterized by their appreciation for the introduction to and the opportunity for engagement in new, meaningful active communities. Of the remaining 21 citizens who were not physically active in civil society organizations before their rehabilitation program, 14 (67%) started and remained active at 6-month follow-up. They recounted receiving suitable support and feeling safe, and attributed their physical activity engagement in civil society organizations to a sense of social belonging and camaraderie. Among the seven citizens who were not active in civil society organizations before or after their rehabilitation program, the support provided was perceived as insufficient to provide safety and meaningfulness considering the substantial health and social issues experienced by these citizens. Compared with the historic control group, 34 percent point fewer citizens were already active in civil society organizations before their rehabilitation program, while 29 percent point more citizens started and remained active in civil society organizations (p=0.004).

**Figure 3:**
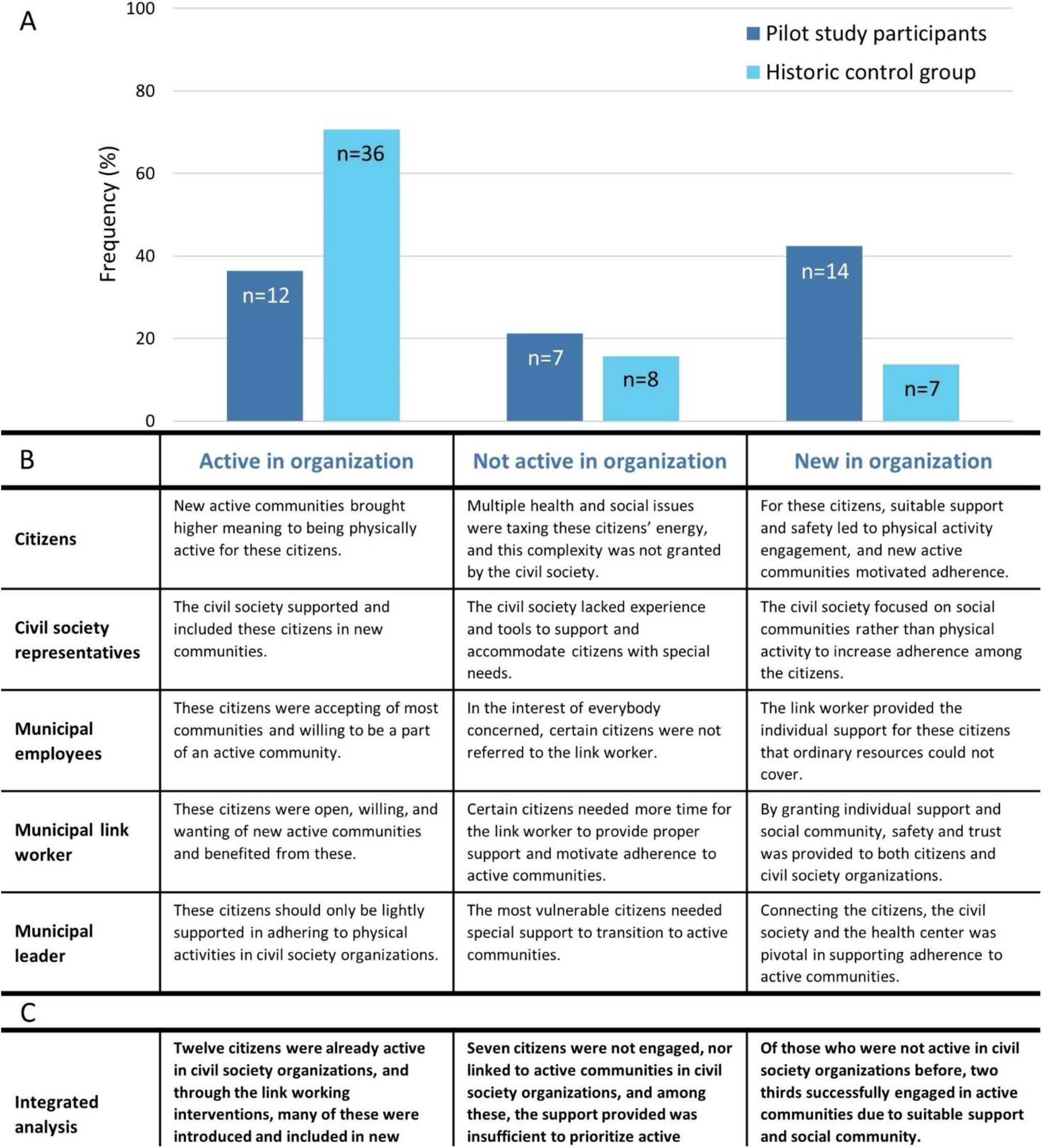
Joint display illustrating A) quantitative classification of participants in the intervention and historic control group into 1. *Active in organization*, 2. *Not active in organization*, and 3. *New in organization*; B) qualitative analysis of focus groups (citizens, civil society representatives, and municipal employees) and individual interviews (municipal link worker and municipal leader); and C) integrated analysis. 1. *Active in organization* includes citizens who were physically active in civil society organizations at baseline and 6-month follow-up; 2. *Not active in organization* includes citizens who were not physically active in civil society organizations at baseline and 6-month follow-up; and 3. *New in organization* includes citizens who were not physically active in civil society organizations at baseline, but physically active in civil society organizations at 6-month follow-up.

General agreement existed among the citizens, civil society representatives, municipal leader and employees, that the link worker succeeded in building trust among the citizens and the civil society organizations, allowing for engagement in shared activities. The presence of social community, e.g., carpooling, social community, and camaraderie, was perceived by the citizens to be highly important for physical activity engagement in civil society organizations. Accordingly, the link worker emphasized changing her focus from prioritizing high physical activity intensities to focusing more on supporting the social community when introducing the citizens to new activities in attempt to improve adherence.

> *In the beginning I believed, they [the citizens] needed to exercise with a certain intensity, and now it’s more about the joy and fun of it, because that’s what makes them come back for more*.

> *(Municipal link worker)*

The citizens generally emphasized that the inspiration and support from the link worker were essential for their engagement in a particular physical activity in a civil society organization. According to the citizens themselves, they needed a push which they welcomed from the link worker. The citizens mostly felt welcome and safe when getting acquainted with new physical activities, either through the visiting program or when visiting civil society organizations on their own.

> *The linking* interventions *have helped because I sometimes need a kick. Otherwise, I wouldn’t have signed up in the organization, I don’t believe so*.

> *(Citizen)*

Still, some citizens described feeling unsafe, misunderstood, and left out, e.g., because of lack of competence and know-how in being physically active in a community, or physical pain and challenges that the civil society organizations could not provide for. Accordingly, the civil society representatives and the link worker described cases of miscommunication of citizens’ physical challenges. As a result, the citizens and civil society representatives concerned, expressed feeling insecure and distrusting, leading to the perception that physiotherapeutic support was required.

Eventually, these citizens felt left on their own to find matching physical activities to engage in.

> *I lost my motivation, when someone from the civil society organization said they didn’t think I fit in because of my bad physical health. But my bad health is exactly why I signed up.\*

> *(Citizen)*

Among the municipal employees there were mixed attitudes about where to place the responsibility for link working. Some employees regarded link working as a crucial part of their work, while others expressed not referring some citizens to the link worker or civil society organizations because they considered them particularly vulnerable and/or beyond reach. They were convinced that it was in the best interest of everybody concerned.

> *If I talk to citizens that has no sense of their own situation or no motivation to be physically active, I am not sure I would recruit them—for the sake of the civil society organization and others*.

> *(Municipal employee)*

### Acceptability of the interventions

#### Link worker

The citizens, civil society representatives, municipal leader, employees, and link worker herself considered it crucial for success that the link worker had a natural ability to foster the relation to 1) the citizens, providing personal support for those in need; and 2) the civil society organizations, learning about their activities, and collaborating in identifying relevant physical activities.

> *We cannot separate the personal and professional qualities. When working in the link of something, it is highly important to be accommodating, listening, and curious—and think, let’s see possibilities instead of limitations. […] It has something to do with the approach in the meeting with others*.

> *(Municipal leader)*

sFigure 1 presents results on familiarity/contact with the link worker and perceived support. Most citizens were familiar with the link worker at baseline, and around half were in contact with the link worker at 3- and 6-month follow-up. Of those who were not in contact with the link worker, 4 of 9, 9 of 15, and 9 of 16 were already physically active at baseline, 3-, and 6-month follow-up, respectively. Two and six months into the study period, respectively, 10 of 10 municipal employees were familiar with the link worker function; 9 of 10 and 10 of 10 municipal employees, respectively, found it relevant for the citizens and their own work; while 5 of 10 and 6 of 10 municipal employees, respectively, had used the function. The civil society representatives, municipal leader, employees, and link worker herself emphasized that the link worker function required anchoring to secure continued individual support for those in need and continued collaboration with civil society organizations. They attributed the success of the linking interventions to the link worker and explained that she managed to create trusting relations to the citizens and sense who needed support. This was attributed to her personality and behavior (i.e., curiosity and openness to change and development), human qualities (i.e., empathy and authenticity) and professional background (i.e., nursing). The municipal employees explained that without a dedicated function, link working would be deprioritized in favor of their main responsibilities.

> *I would love to say that without the link worker, we would continue in the same way. But it is not going to happen—it will drown in our main work responsibilities*.

> *(Municipal* employee)

The link worker was responsible for initializing and managing the remaining linking interventions (i.e., the visiting program, co-created activities, and the digital platform).

#### Visiting program and co-created activities

The municipal employees expressed that due to the integration of the visiting program into the municipal rehabilitation program and the presence of the link worker, the citizens seemed more open and willing to experience different physical activities in new communities.

> *In my experience, the most important component in the linking interventions, is that there is someone dedicated to following and introducing the citizens to activities in civil society […]*.

> *(Municipal employee)*

At baseline, 20 of 30 citizens had experience with the visiting program, and at 3- and 6-month follow-up, 16 of 29 and 18 of 30, respectively, had experience with physical activities in civil society organizations during the preceding 3 months. sFigure 1 presents results on satisfaction with the visiting program. Reasons for having no experience included lack of motivation, not feeling safe, not finding the level suitable, and not appreciating the community; each of these were reported by ≤2 participants at 3- and 6-month follow-up, respectively. Other reasons included already engaging in physical activities in a civil society organization (6 of 12 and 4 of 12, respectively) and/or on their own (7 of 12 and 8 of 12, respectively); or not being interested in engaging in physical activities (2 of 12 and 2 of 12, respectively) at 3- and 6-month follow-up. The visiting program included collaboration with 10 civil society organizations, and several citizens voiced a wish for experiencing even more physical activities to inspire and match their needs. The visiting program was not tested in cardiac rehabilitation programs, although the citizens participating expressed curiosity and a wish to experience different physical activities in civil society organizations.

> *Naturally, it would be easier to get out and try something new if we were going as a team knowing each other well—that’s for certain*.

> *(Citizen)*

Only few physical activities were co-created in the local community; however, civil society representatives expressed a wish to engage in collaboration with others to provide active communities for citizens to be more physically active.

> *I’ve always believed that as long as the citizens become active, then I don’t care whether it is walking, strength training or walking soccer. Just get off the couch*.

> *(Civil society representative)*

#### Digital platform

The municipal leader, employees, and link worker considered the digital platform a useful tool for the link worker and other municipal employees to support citizens in transitioning to physical activities in civil society. Moreover, the link worker expressed that the platform was a relevant tool in the dialogue and collaboration with civil society organizations.

> *Maybe their [the civil society organizations’] activities won’t be used in the first while, but when the activities are uploaded to the digital platform, they know that I see them and acknowledge them and our collaboration*.

> *(Municipal link worker)*

At baseline, 18 of 32 citizens were familiar with the digital platform, while 8 of 11 and 8 of 10 of the municipal employees had experience with the platform two and six months into the study period, respectively. Of these, 7 of 8 and 6 of 8 had used the platform; 6 of 8 and 5 of 8 had found relevant activities for their citizens; and 8 of 8 and 8 of 8 found that the platform was useful in their work with the citizens at two and six months, respectively. The municipal employees and link worker considered continued success to be dependent on future possibilities of keeping the platform updated, and further development to accommodate interactivity and upscaling of the linking interventions.

### Citizens’ physical and mental health

Changes in citizens’ physical and mental health from baseline to 3- and 6-month follow-up, respectively, are presented in Figure 4. From baseline to 6-month follow-up LPA time increased by a mean (95% CI) of 15.4 (2.3; 28.5) min/day, while sitting time increased by 39.6 (9.8; 69.3) min/day from baseline to 3-month follow-up. All other outcomes remained unchanged from baseline to 3- and 6-month follow-up. sFigure 2 presents differences from pre-baseline to baseline and 6-month follow-up, respectively. From pre-baseline to baseline (i.e., during the rehabilitation program), steps and total physical activity decreased, 6-minute walk distance increased, and all other outcomes remained unchanged. From pre-baseline to 6-month follow-up, muscle mass and body weight decreased, 6-minute walk distance increased, and all other outcomes remained unchanged.

**Figure 4:**
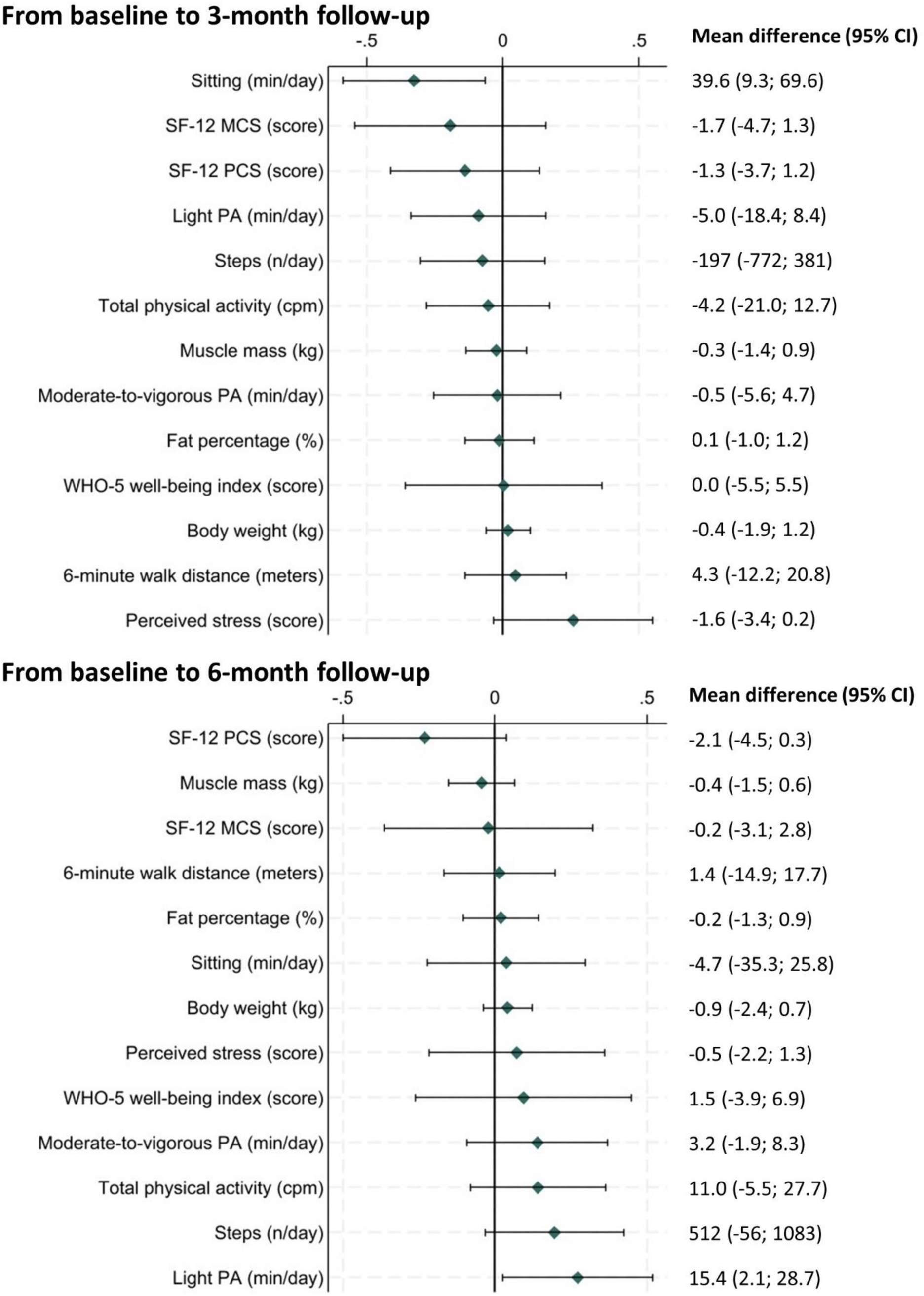
Standardized mean differences and least square mean differences, and associated 95% confidence intervals from baseline to 3-month follow-up; and baseline to 6-month follow-up. Data include objectively measured moderate-to-vigorous physical activity time; light physical activity time; sitting time; total physical activity level; steps; 6-minute walk distance; body weight; fat percentage; muscle mass; SF-12 physical and mental component summary; WHO-5 well-being index; and perceived stress. Standardized mean differences are reversed for sitting time; body weight; fat percentage; and perceived stress. SF-12, 12-item short-form health survey; MCS, mental component summary; PCS, physical component summary; PA, physical activity; WHO, World Health Organization.

No changes were observed in Tandem balance score, social isolation, or severe loneliness, neither from baseline to 3- and 6-month follow-up, nor from pre-baseline to baseline and 6-month follow-up.

### Collaboration between the municipality and civil society organizations

The municipal leader, employees, link worker, and civil society representatives explained that before project Active Communities was initiated, the collaboration between the municipality and the civil society organizations had slowly started but had come to a halt. They expressed that the co-creation process facilitated productive collaboration, mutual understanding, and appealed all parties to be curious and constructive. According to them, this was possible despite great disagreements in the past and prejudice that responsibilities were imposed on to one another. In the end, they all acknowledged that everyone had a joint wish for strengthening the link between the municipality and the civil society. According to the municipal leader, the collaboration would not have been possible if the municipality had initiated the linking interventions on their own initiative. She explained that building on a partnership and inviting all actors to join the process of co-creating the linking interventions, gave the link worker a mandate to test new initiatives in the civil society.

> *I think, the way* that *we approached this has given a greater mandate for change compared to hiring a link worker on my own initiative […] we have actually worked professionally to generate the knowledge which is now implemented*.

> *(Municipal leader)*

Despite the civil society representatives expressing a generally positive view of the co-creation and collaboration processes, only a few civil society organizations were attentive, willing, and able to create new classes accommodating the needs of the target group and/or actively engage in securing a safe welcoming into existing classes.

According to one of the municipal employees, the visibility of the municipal link worker and her engagement in productive collaboration with the civil society organizations positively affected the civil society organizations’ openness to engage in collaboration with other departments in the municipality. However, civil society representatives expressed that if the municipality failed to anchor the link worker function, this would be perceived as belittling of their work.

> *If all this work falls* apart*, it is yet another proof that the work of volunteers in civil society organizations is not recognized*.

> *(Civil society representative)*

### Organizational development

The municipal leader, employees, and link worker expressed that the involvement in project Active Communities led to a greater focus on link working in the municipality. They mentioned that this involved a change in procedures in the health center to include link working as a referring option in the citizen records. The health professionals expressed a professional satisfaction in having a colleague specialized in link working to whom they can refer their citizens, and thereby ensure that the support they themselves have provided was not in vain.

> *The link worker helps* the *citizens after their rehabilitation program. It is very satisfying for me as a therapist, knowing that she helps them if they have the need*.

> *(Municipal employee)*

Yet, the responsible team for cardiac rehabilitation programs did not find a solution to fully integrate link working in these programs. The team argued that the reason for this was legal obligations to ensure rehabilitation including proper intensity and type of physical activities. They felt unsure that these obligations could be fulfilled, considering uncertainties regarding the exercise equipment in the civil society organizations and their own competencies to potentially provide alternative rehabilitation programs. However, they mentioned that they started arranging staff meetings in different civil society organization to explore the possibilities of referring their citizens to continued physical activity after their rehabilitation program ends.

The link worker explained having to be very insistent for her colleagues to change their work procedures, and the municipal leader expressed astonishment over her employees’ insecurity and resistance towards change.

> *Some of my employees had a hard time moving the programs out into the civil society organizations. […] We had a lot of talks about very practical matters. […] I was surprised that it was that hard for them to move out of the health center*.

> *(Municipal leader)*

The municipal leader trusted that the involvement of several stakeholders in a co-creation process gave her a mandate to insist on effectuating the commonly decided linking interventions. The municipal leader emphasized that the link worker function is highly exposed to economization in the municipality, and that focusing on documenting and structuring the initiatives will be important for future retainment of the function and the overall development in the municipality.

## Discussion

The main finding of this study was that 67% of the citizens who were not physically active in civil society organizations before their rehabilitation program, started and remained active at 6-month follow-up—significantly more than in the historic control group. In line with this study, previous studies have found that suitable support and social community are key elements for a strong transitioning, and contributing to this are key elements such as general sustainability, weekly regularity, and easy accessibility (2, 5, 25). The positive effect on physical activity engagement is most likely closely linked to the high acceptability of the linking interventions among all participants involved.

Further, we observed an increase in LPA time over the 6 months following the rehabilitation program. This was accompanied by insignificant increases in MVPA time, TPA, and step number— overall pointing to a general positive influence on physical activity level. This is supported by previous reviews reporting general increases in physical activity following rehabilitation programs with initiatives for stronger transitioning from rehabilitation to civil society organizations (2, 26).

Although speculative, these findings may indicate a mitigation of the general decrease in physical activity level occurring with aging (27) and among persons with chronic conditions (16).

Among the identified challenges in linking a municipal health center with civil society organizations, we found that for some of the previously inactive citizens, the support provided was insufficient to fulfill their needs and allow for personal issues related to health and social circumstances. Although the interventions were tailored to the target group by involving them in the co-creation process, the most vulnerable citizens were not successfully represented (7), potentially explaining why the needs of these citizens were not sufficiently matched. This unmatching of needs entailed that these citizens were potentially confirmed in their experience of feeling mistaken when attempting to take up a new active habit, and thus resigning themselves further on the possibilities of being physically active in a civil society organization. The failure to match these citizens’ needs may additionally result from the miscommunication related to these citizens’ physical challenges between the link worker and the civil society organizations. This may further have resulted in limited adaptation and creation of classes to accommodate these citizens’ needs. Indeed, a previous paper found that shifting of traditional sport culture was one of nine challenges involved in developing new physical activity opportunities (11)—an aspect that were not in focus nor provided for in this project. Involving social community groups and organizations from civil society in the collaboration can potentially provide the support needed for these citizens to engage in civil society activities. Moreover, challenges included lack of integration of the co-created linking interventions into cardiac rehabilitation programs, mainly explained by perceived barriers of legal obligations regarding rehabilitation content. Evaluation of these perceived barriers by an expert in municipal legal obligations, supported by a larger focus on incorporating the linking interventions into the health professionals’ practice, would potentially have accommodated this.

Besides miscommunication related to citizens’ physical challenges, the collaboration between the municipality and civil society organizations were characterized by a mutual understanding and mission. This study supports previous findings that lack of medical knowledge in civil society organizations, lack of time among health professionals, and different interests and cultures are barriers that should be considered when aiming to strengthen the collaboration between a public health institution and civil society organizations (10). In the present study, lack of time among health professionals were provided against by employing a dedicated link worker, while different interests and cultures were addressed in the co-creation process (7). This included aspects of unmatching of expectations and former disagreements—however, primarily dominating the initial collaboration. The fact that some citizens felt mistaken partly due to their health issues, and that the link worker and civil society representatives perceived physiotherapeutic support a requirement, indicate that lack of medical knowledge in civil society organizations were indeed perceived as an essential barrier for collaboration and link working.

Project Active Communities led to a political reprioritization of economic resources in Odsherred municipality in 2023 to support future link working between the municipality and civil society organizations (28). Social prescribing including linking citizens to physical activities in civil society is increasingly advocated by the Danish Health Authority, particularly among citizens with chronic conditions, but with no specific recommendations for practical implementation (29). This may be attributed to the sparse evidence in the area (9). Yet, in 2020 almost half of 40 responding Danish municipalities performed some kind of structured link working with varying elements (30) and using different models for social prescribing (30, 31). Social prescribing was developed in England and implemented in primary care across the country (8, 32) showing general health benefits (33), and has been adopted by several other countries to fit their specific contexts (32).

Limitations of this study include the lack of a true concurrent control group for comparison, challenging the interpretation of results and inferences to whether a general age-related decrease in physical activity level have been mitigated as previously discussed. Moreover, the low number of participants recruited in the intervention study resulted in uncertain estimates (i.e., wide CIs) of the secondary outcomes. As the recruitment rate was rather high, corresponding to 63.5%, the low participant number was due to few citizen referrals and low participation in municipal rehabilitation programs. To investigate the representativeness of the citizens enrolled in a municipal rehabilitation program, questionnaire data were collected from a separate reference group (n=52) shortly before the linking interventions were implemented in the municipal health center. These data indicate that more citizens in the reference group were already engaged in physical activities in a civil society organization, and that they were generally more physically active compared with the intervention group participants (sTable 2). Further, the representativeness of the general population in the municipality was likely limited, as referral to municipal rehabilitation programs seem to be dependent on citizens’ socioeconomic status (34). We did not collect data on ethnicity, but observations during data collection indicate that the ethnic representation was highly limited. Altogether, the study sample may not represent the most vulnerable citizens in the general population, but among those engaged in rehabilitation programs, the citizens with the greatest need for support appear to be represented. This representativeness was carried over in the qualitative data collection related to the citizens, as these participants were sampled from the intervention group.

However, the additional representation of civil society organizations, municipal employees, leader, and link worker in the qualitative data collection ensured representation of multiple perspectives and contributed to achieving information power. Another limitation of this study was the assessment method used to evaluate standing balance. Most participants obtained maximal scores at all time points, while few obtained sub-maximal scores at pre-baseline and maximal scores at subsequent time points—however, this improvement was not detectable in the analysis. Still, all assessment methods were perceived well by most intervention group participants (sFigure 3).

Everything considered, the mixed methods design including triangulation of methods, sources, and researchers is a major strength of this study. This involved separate quantitative and qualitative analysis with subsequent integration for a comprehensive understanding of the link between the municipality and civil society organizations, and how this affected physical activity engagement among citizens.

## Conclusions

In conclusion, we observed an increase in physical activity engagement in civil society organizations following co-created interventions to link a municipal health center and civil society organizations. This was assisted by experienced high acceptability of the linking interventions among participants, an increase in citizens’ LPA time, and strengthened collaboration between the municipal health center and civil society organizations. This study extends the sparse evidence for social prescribing, suggesting co-creation of interventions as a method to link municipal rehabilitation programs with physical activities in civil society organizations. Future studies should investigate whether this is transferable to other target groups, other municipalities, and other health settings.

## Data Availability

The datasets used and/or analyzed during the current study are available from the corresponding author on reasonable request.

## List of abbreviations

BMI: body mass index
CI: confidence interval
CPM: counts per minute
CVD: cardiovascular diseases
ISCED-2011: International Standard Classification of Education 2011
LPA: light physical activity
MMR-RHS: Mixed Methods Reporting in Rehabilitation & Health Sciences
MVPA: moderate-and-vigorous physical activity
PSS: the perceived stress scale
SF-12: the 12-item short-from health survey
T2D: type 2 diabetes
T-ILS: the three-item loneliness index
TPA: total physical activity
WHO: World Health Organization
WHO-5: the 5-item
WHO: well-being index

## Declarations

### Ethics approval and consent to participate

According to the Scientific Ethical Committee for The Capital Region of Denmark, this study did not require ethical approval (reference-no.: 21076311). Prior to participation, informed consent was obtained from all the participating persons. The study was conducted in accordance with the Data Protection Regulation and complies with the General Data Protection Regulation (GDPR) (EU) 2016/679.

### Consent for publication

Not applicable.

## Authors’ contributions

IKT, MLH, KT, HS, HOJ, TPA, JKL, ABGH, and MR-L contributed to the design of the study. IKT, MLH, KT, and MR-L planned and conducted the data collection. IKT, JM, MLH, and MR-L analyzed and interpreted the data. With support from JM, MLH and MR-L, IKT wrote the first draft of the manuscript. All authors read and approved the final manuscript.

## Acknowledgements

We acknowledge all participants in the study—citizens, civil society representatives, municipal employees, the municipal leader, and link worker—and the partnering research institutions, as well as the funds that supported this study. The Centre for Physical Activity Research (CFAS) is supported by TrygFonden (Grants ID 101390, ID 20045, and ID 125132).

## Literature

1. Pedersen BK, Saltin B. Exercise as medicine - evidence for prescribing exercise as therapy in 26 different chronic diseases. Scand J Med Sci Sports. 2015;25 Suppl 3:1–72.

2. Thomsen S, Kristensen GDW, Jensen NWH, Agergaard S. Maintaining changes in physical activity among type 2 diabetics - A systematic review of rehabilitation interventions. Scand J Med Sci Sports. 2021;31(8):1582–91.

3. Umpierre D, Ribeiro PA, Kramer CK, Leitao CB, Zucatti AT, Azevedo MJ, et al. Physical activity advice only or structured exercise training and association with HbA1c levels in type 2 diabetes: a systematic review and meta-analysis. JAMA. 2011;305(17):1790–9.

4. Thorsen IK, Kayser L, Teglgaard Lyk-Jensen H, Rossen S, Ried-Larsen M, Midtgaard J. “I Tried Forcing Myself to do It, but Then It Becomes a Boring Chore“: Understanding (dis)engagement in Physical Activity Among Individuals With Type 2 Diabetes Using a Practice Theory Approach. Qual Health Res. 2022;32(3):520–30.

5. Engdal S, Hansen HF, Ottesen LS. Mind the gap: building bridges between public sector exercise programmes and civil society sports associations. An integrative review of the literature. European Journal for Sport and Society. 2023;20(3):279–98.

6. VIVE The Danish Center for Social Science Research. Frivillighedsundersøgelse 2020 - En repræsentative befolkningsundersøgelse af udviklingen i danskernes frivillige arbejde [Voluntariness study 2020 - A representative population survey of the development in Danish citizens’ voluntary work] 2021 [Accessed 2024 24 Jun]. Available from: https://www.vive.dk/media/pure/lxmk93vd/5662919.

7. Hansen ABG, Hansen ML, Golubovic S, Bloch P, Lorenzen JK, Almdal TP, et al. Co-creating active communities: processes and outcomes of linking public rehabilitation programs with civic engagement for active living in a Danish municipality. Res Involv Engagem. 2023;9(1):83.

8. National Health Service (NHS) England, NHS Improvement. Social prescribing and community-based support 2020 [Accessed 2024 17 Jun]. Available from: https://www.england.nhs.uk/wp-content/uploads/2020/06/social-prescribing-summary-guide-updated-june-20.pdf.

9. Midtgaard J, Christensen JR, MacDonald CS, Færch M. Social prescribing in combating loneliness and supporting physical activity. Ugeskr Laeger. 2023;185(11).

10. Leenaars KE, Smit E, Wagemakers A, Molleman GR, Koelen MA. Facilitators and barriers in the collaboration between the primary care and the sport sector in order to promote physical activity: A systematic literature review. Prev Med. 2015;81:460–78.

11. Staley K, Donaldson A, Randle E, Nicholson M, O’Halloran P, Nelson R, et al. Challenges for sport organisations developing and delivering non-traditional social sport products for insufficiently active populations. Aust N Z J Public Health. 2019;43(4):373–81.

12. Fetters MD, Curry LA, Creswell JW. Achieving integration in mixed methods designs-principles and practices. Health Serv Res. 2013;48(6 Pt 2):2134–56.

13. DinGeo. Nøgletal for Odsherred Kommune [Key figures for Odsherred Municipality] 2022 [Accessed 2024 17 Jun]. Available from: https://www.dingeo.dk/kommune/odsherred/.

14. Poulsen HS, Eiriksson SD, Chistiansen ASJ, Wingstrand A. Sundhedsprofil 2021 for Region Sjælland og kommuner - “Hvordan har du det?” [Health profile 2021 for Region Zealand and municipalities - “How are you?”]. Region Sjælland, Data og udviklingsstøtte; 2022.

15. Levitt HM, Bamberg M, Creswell JW, Frost DM, Josselson R, Suarez-Orozco C. Journal article reporting standards for qualitative primary, qualitative meta-analytic, and mixed methods research in psychology: The APA Publications and Communications Board task force report. Am Psychol. 2018;73(1):26–46.

16. Thorsen IK, Yang Y, Valentiner LS, Glumer C, Karstoft K, Brond JC, et al. The Effects of a Lifestyle Intervention Supported by the InterWalk Smartphone App on Increasing Physical Activity Among Persons With Type 2 Diabetes: Parallel-Group, Randomized Trial. JMIR Mhealth Uhealth. 2022;10(9):e30602.

17. A. T. S. Committee on Proficiency Standards for Clinical Pulmonary Function Laboratories. ATS statement: guidelines for the six-minute walk test. Am J Respir Crit Care Med. 2002;166(1):111–7.

18. Guralnik JM, Simonsick EM, Ferrucci L, Glynn RJ, Berkman LF, Blazer DG, et al. A short physical performance battery assessing lower extremity function: association with self-reported disability and prediction of mortality and nursing home admission. J Gerontol. 1994;49(2):M85–94.

19. Ware J, Jr., Kosinski M, Keller SD. A 12-Item Short-Form Health Survey: construction of scales and preliminary tests of reliability and validity. Med Care. 1996;34(3):220–33.

20. World Health Organization. Regional Office for Europe. Wellbeing measures in primary health care: the DepCare project : report on a WHO meeting Stockholm, Sweden 12-13 February 1998: WHO Regional Office for Europe; 1998.

21. Cohen S, Kamarck T, Mermelstein R. A global measure of perceived stress. J Health Soc Behav. 1983;24(4):385–96.

22. Valtorta NK, Kanaan M, Gilbody S, Hanratty B. Loneliness, social isolation and risk of cardiovascular disease in the English Longitudinal Study of Ageing. Eur J Prev Cardiol. 2018;25(13):1387–96.

23. Hughes ME, Waite LJ, Hawkley LC, Cacioppo JT. A Short Scale for Measuring Loneliness in Large Surveys: Results From Two Population-Based Studies. Res Aging. 2004;26(6):655–72.

24. Malterud K, Siersma V, Guassora AD. Information power: Sample content and size in qualitative studies. Qualitative research in psychology: Expanding perspectives in methodology and design, 2nd ed. Washington, DC, US: American Psychological Association; 2021. p. 67–81.

25. Albarracín D, Fayaz-Farkhad B, Granados Samayoa JA. Determinants of behaviour and their efficacy as targets of behavioural change interventions. Nature Reviews Psychology. 2024.

26. Onerup A, Arvidsson D, Blomqvist A, Daxberg EL, Jivegard L, Jonsdottir IH, et al. Physical activity on prescription in accordance with the Swedish model increases physical activity: a systematic review. Br J Sports Med. 2019;53(6):383–8.

27. Danish Health Authority, University of Southern Denmark. Opfylder ikke WHOs anbefalinger for fysisk aktivitet [Do not meet WHO recommendations on physical activity] 2023 [Accessed 2024 30 May]. Available from: https://proxy.danskernessundhed.dk/SASVisualAnalyticsViewer/VisualAnalyticsViewer_guest.jsp?reportName=Opfylder%20ikke%20WHOs%20anbefalinger%20for%20fysisk%20aktivitet&reportPath=/Produktion/Danskernes_Sundhed/&reportViewOnly=true.

28. Trivsels-Sundheds-og Forebyggelsesudvalget - Odsherred Kommune [Welfare Health and Prevention Committee - Odsherred Municipality]. Referat 16. januar 2024 kl. 08.30 [Minutes January 16, 2024, at 8:30 AM] 2024 [Accessed 2024 17 Jun]. Available from: https://dagsordener.odsherred.dk/vis?Referat-Trivsels-%2C-Sundheds--og-Forebyggelsesudvalget-d.16-01-2024-kl.08.30&id=00e5e6fb-9dbd-40db-a6b4-2a9acd428a4d.

29. Danish Health Authority. Forebyggelsestilbud til borgere med kronisk sygdom - Kvalitetsstandarder [Prevention offers for citizens with chronic conditions - Quality standards]. Copenhagen2024.

30. Engdal Larsen S, Ottesen LS, Hansen HF. Implementering af brobygning: Fra kommunalt træningsforløb til idrætsforeninger eller idrætsfællesskaber [Implementation of bridge-building: From municipal exercise programmes to voluntary sports associations or sports communities]. Center for Holdspil og Sundhed, Institut for Idræt og Ernæring; 2020.

31. Husk K, Blockley K, Lovell R, Bethel A, Lang I, Byng R, et al. What approaches to social prescribing work, for whom, and in what circumstances? A realist review. Health Soc Care Community. 2020;28(2):309–24.

32. Morse DF, Sandhu S, Mulligan K, Tierney S, Polley M, Chiva Giurca B, et al. Global developments in social prescribing. BMJ Glob Health. 2022;7(5).

33. Chatterjee HJ, Camic PM, Lockyer B, Thomson LJM. Non-clinical community interventions: a systematised review of social prescribing schemes. Arts & Health. 2018;10(2):97–123.

34. Sortso C, Lauridsen J, Emneus M, Green A, Jensen PB. Socioeconomic inequality of diabetes patients’ health care utilization in Denmark. Health Econ Rev. 2017;7(1):21.

