## Supplementary material for "Effect and acceptability of co-created interventions linking public rehabilitation programs with civil society involvement for physical activity engagement – a convergent mixed methods pilot study"

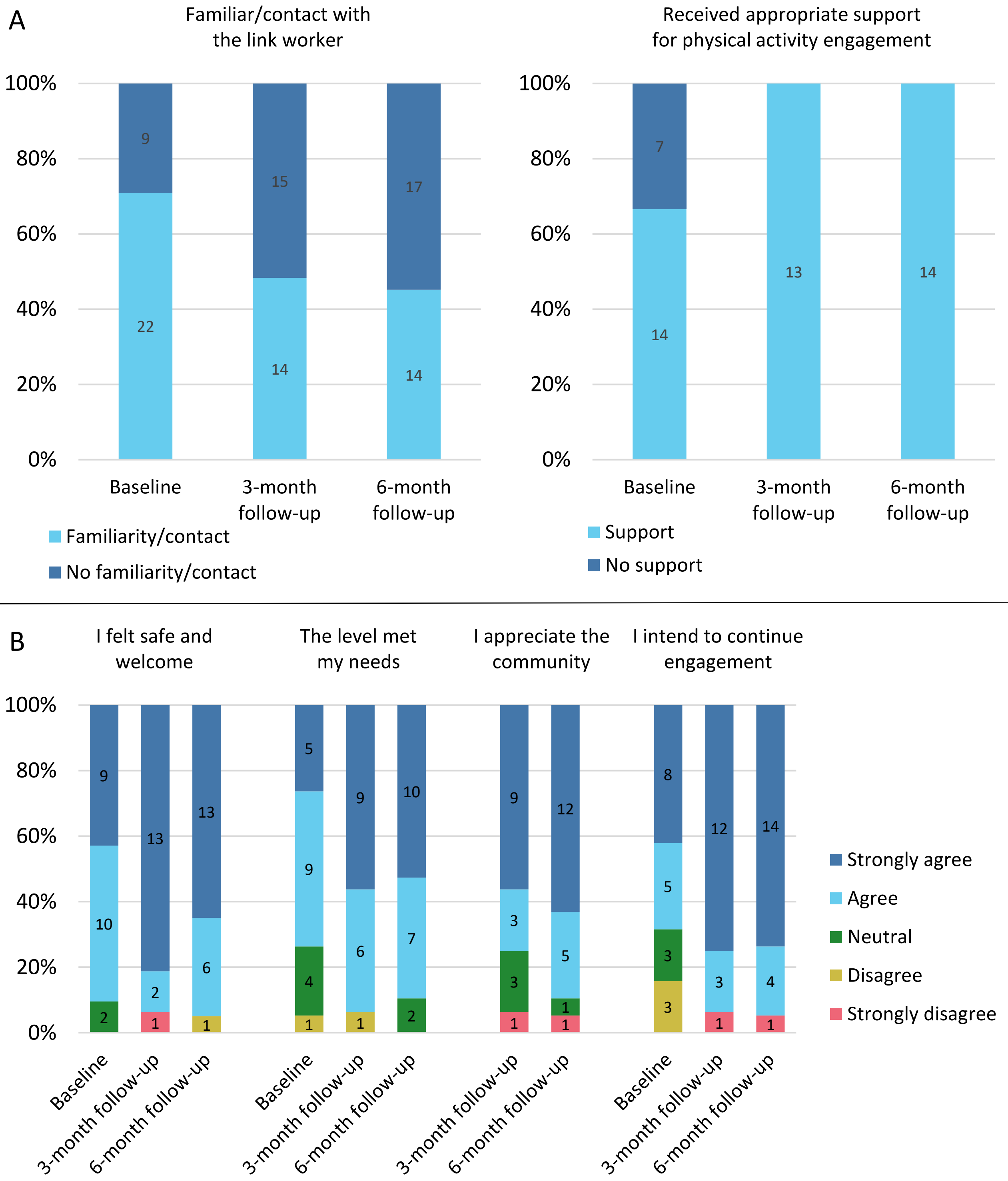


**sFigure 1:** A) Familiarity/contact and perceived support from the link worker, and B) satisfaction with the visiting program at Baseline, 3- and 6-month follow-up.

**From pre-baseline to baseline**

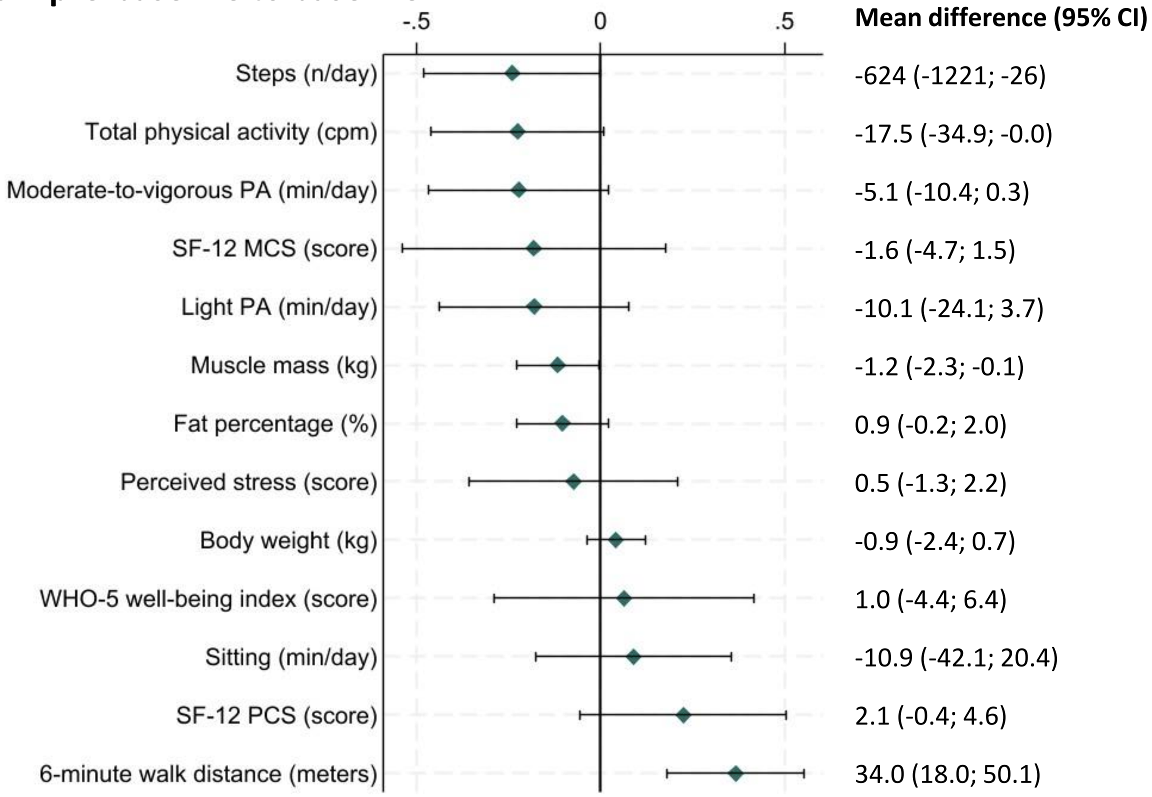


**From pre-baseline to 6-month follow-up**

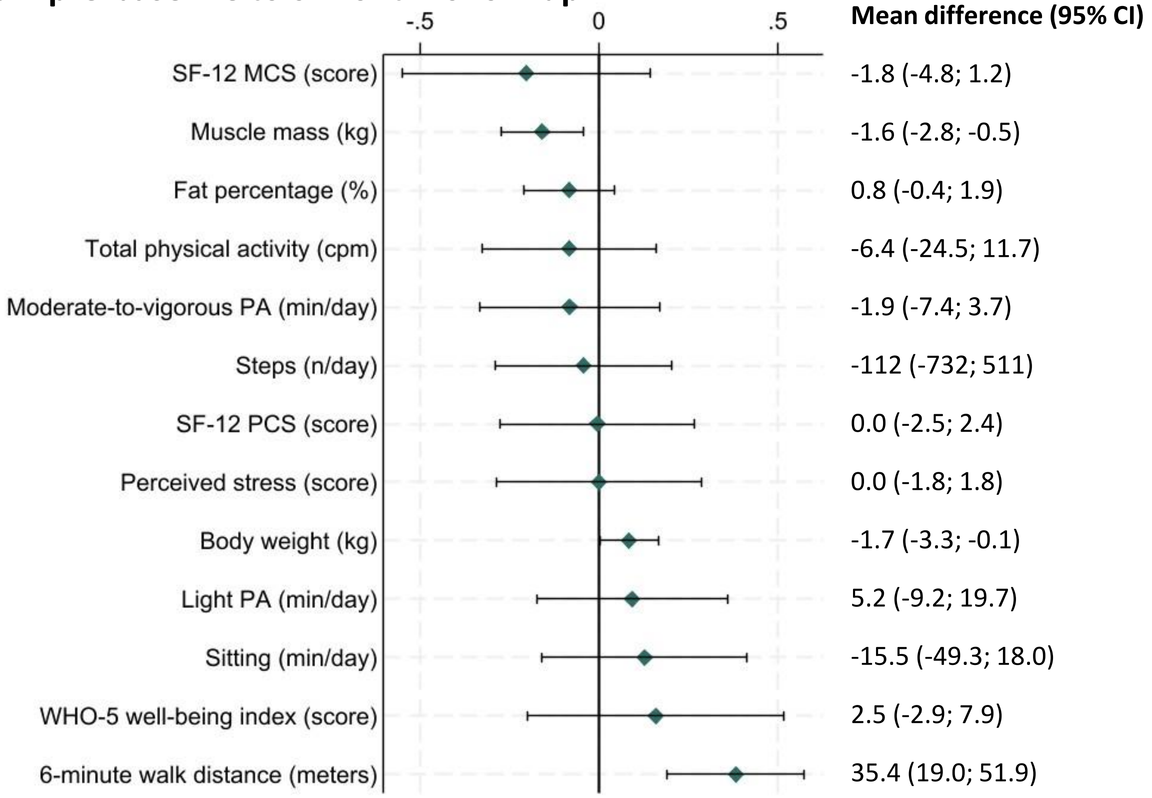


**sFigure 2:** Standardized mean differences and least square mean differences, and associated 95% confidence intervals from pre-baseline to baseline; and pre-baseline to 6-month follow-up. Data include objectively measured moderate-to-vigorous physical activity time; light physical activity time; sitting time; total physical activity level; steps; 6-minute walk distance; body weight; fat percentage; muscle mass; SF-12 physical and mental component summary; WHO-5 well-being index; and perceived stress. Standardized mean differences are reversed for sitting time; body weight; fat percentage; and perceived stress. SF-12, 12-item short-form health survey; MCS, mental component summary; PCS, physical component summary; PA, physical activity; WHO, World Health Organization.

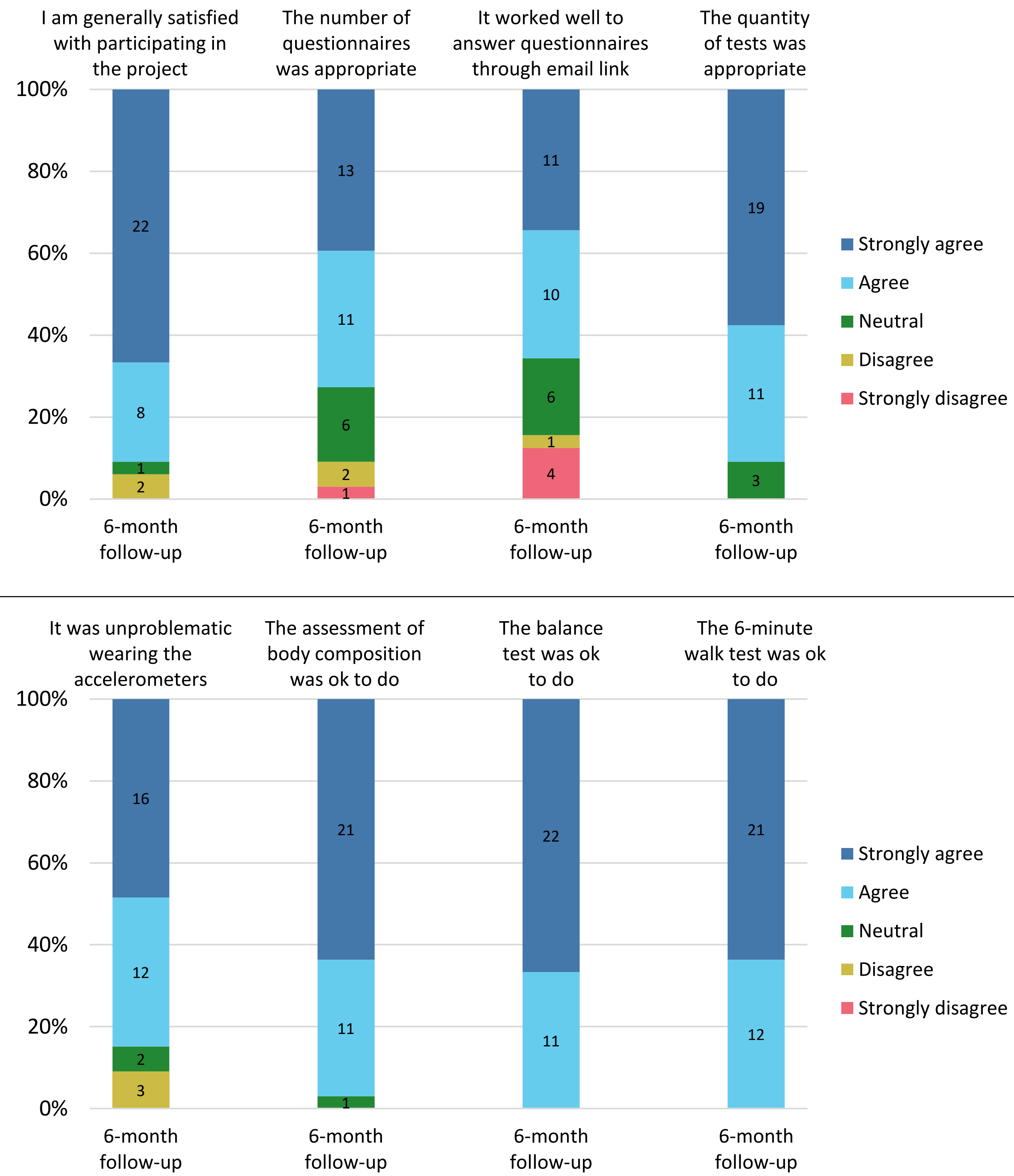


**sFigure 3:** Satisfaction related to intervention study participation and assessment methods, including questionnaires, physical tests, and objective measurement of physical activity.

**sTable 1:** Quantitative outcomes, qualitative themes and reporting approach for integration.

| Results subsections | Program theory elements | Quantitative outcomes | Qualitative themes | Reporting approach for integration |
|---|---|---|---|---|
| 1. Participant characteristics | N/A | Participant recruitment (Citizens, Civil society representatives, Municipal employees)<br>Sociodemographic and disease-related characteristics (Citizens) | N/A | N/A, Quantitative only |
| 2. Physical activity engagement | Citizen's engagement in physical activity | Categorization of physical activity engagement in civil society organizations based on monthly reporting (Citizens) | Physical activity engagement | Joint display<br>(Figure 2) |
| 3. Acceptability of the interventions: *Link worker* | Link worker | Familiarity/contact (Citizens)<br>Appropriate support (Citizens)<br>Familiarity (Municipal employees)<br>Applicability (Municipal employees)<br>Utilization (Municipal employees) | | Narrative |
| 3. Acceptability of the interventions: *Visiting program and co-created activities* | Visiting program and co-created activities | Experience (Citizens)<br>Safety (Citizens)<br>Suitability (Citizens)<br>Community (Citizens)<br>Potential (Citizens) | Acceptability of the interventions | Narrative |
| 3. Acceptability of the interventions: *Digital platform* | Digital platform | Familiarity (Citizens)<br>Experience (Municipal employees)<br>Utilization (Municipal employees)<br>Applicability (Municipal employees) | | Narrative |
| 4. Citizens' physical and mental health | Citizens' physical and mental health | Moderate-and-vigorous physical activity time (Citizens)<br>Light physical activity time (Citizens)<br>Sitting time (Citizens)<br>Total physical activity (Citizens)<br>Steps (Citizens)<br>6-minute walk distance (Citizens)<br>Tandem score (Citizens)<br>Body weight (Citizens)<br>Fat percentage (Citizens)<br>Muscle mass (Citizens)<br>Body mass index (BMI) (Citizens)<br>Physical health-related quality of life score (Citizens)<br>Mental health-related quality of life score (Citizens)<br>Mental well-being score (Citizens)<br>Perceived stress score (Citizens) | N/A | N/A, Quantitative only<br>(Figure 3) |
| 5. Collaboration between the municipality and civil society | Collaboration between the municipality and civil society | N/A | Collaboration between the municipality and the civil society | N/A, Qualitative only |
| 6. Organizational development | Organizational development | N/A | Organizational development | N/A, Qualitative only |

Overview of results subsections, program theory elements, quantitative outcomes, qualitative themes, and reporting approaches for integration. N/A, not applicable.

**sTable 2:** Characteristics of pilot study participants and the reference group

| | **Pilot study participants** | **Reference group** |
|---|---|---|
| **n** | 33 | 56 |
| **Sex** | | |
| Women | 19 (57.6) | 34 (60.7) |
| Men | 14 (42.4) | 22 (39.3) |
| **Age (years)** | 67.6 (63.9; 74.1) | 63 (52; 70) |
| **Disease group** | | |
| Type 2 diabetes | 6 (18.2) | 15 (28.3) |
| Cardiovascular disease | 9 (27.3) | 13 (24.5) |
| Overweight | 18 (54.5) | 25 (47.2) |
| **Already engaged in physical activities in a civil society organization** | | |
| Yes | 12 (36.4) | 13 (23.6) |
| No | 21 (63.6) | 42 (76.4) |
| **Physical activity level scale (Saltin-Grimpy)** | | |
| Physically inactive | 9 (27.3) | 9 (16.4) |
| Some light physical activity | 18 (54.5) | 37 (67.3) |
| Regular physical activity and training | 6 (18.2) | 8 (14.5) |
| Regular hard physical training for competitive sports | 0 (0.0) | 1 (1.8) |
| **Wish to be more physically active** | | |
| Yes | 31 (93.9) | 48 (87.3) |
| No | 0 (0.0) | 4 (7.3) |
| Maybe | 2 (6.1) | 3 (5.4) |
| **Physical activity level** | | |
| Inactive | 10 (30.3) | 16 (29.1) |
| Inadequate | 17 (51.5) | 24 (43.6) |
| Adequate | 4 (2.1) | 7 (12.7) |
| Optimal | 2 (6.1) | 8 (14.6) |

Data are presented as n (%) and median ($25^{th}$; $75^{th}$ quartile).

### Appendix 1: Acceptability of the interventions

*Link worker*

| | |
|---|---|
| **Citizens** | |
| Familiarity | |
| Baseline | "[Name] is employed in the municipality as a link worker between the municipal programs and the local physical activities in the civil society. Do you know who [name] is? |
| 3-month follow-up | "[Name] is employed in the municipality as a link worker between the municipal programs and the local physical activities in the civil society. Have you been in contact with [name] after your municipal program ended?" |
| 6-month follow-up | "[Name] is employed in the municipality as a link worker between the municipal programs and the local physical activities in the civil society. Have you been in contact with [name] during the last three months?" |
| Suitability | |
| Baseline | "If yes, did you receive support from [name] in finding a relevant physical activity offer?" |
| 3-month follow-up | "If yes, did you receive the necessary support from [name] after your municipal program ended?" |
| 6-month follow-up | "If yes, did you receive the necessary support from [name] during the last three months?" |
| **Municipal employees** | |
| Familiarity | |
| Two and six months into the study period | "[Name] is employed in the municipality as a link worker between the municipal programs and the local physical activities in the civil society. Are you familiar with [Name]'s function?" |
| Relevance | |
| Two and six months into the study period | "If yes, do you find the function relevant for the citizens?" |
| Two and six months into the study period | "If yes, do you find the function relevant for your work?" |
| Use | |
| Two and six months into the study period | "If yes, have you used the link worker function in your work with the citizens (e.g., referred a citizen to [Name] or discussed with [Name] about relevant physical activity offers for a citizen)?" |

*Visiting program and co-created activities*

| **Citizens** | |
|---|---|
| Familiarity | |
| Baseline | "In connection to your municipal program, have you tried one or several physical activities in an organization or similar?" |
| 3-month follow-up | "Have you tried one or several physical activities in an organization or similar after your municipal program ended?" |
| 6-month follow-up | "Have you tried one or several physical activities in an organization or similar during the last three months?" |
| Perception – "I felt safe and welcome" | |
| Baseline | "I was presented with one or several organizations that gave me a good reception and made me feel safe." |
| 3- and 6-month follow-up | "I feel safe and welcome in the physical activity offer." |
| Perception – "The level met my needs" | |
| Baseline | "I engaged in one or several physical activities in civil society organizations that fit my needs." |
| 3- and 6-month follow-up | "The level in the physical activity offer fit my needs." |
| Perception – "I appreciate the community" | |
| 3- and 6-month follow-up | "The community in the physical activity offer is good." |
| Perception – "I intend to continue engagement" | |
| Baseline and 3-month follow-up | "I can imagine continuing this physical activity in civil society, or the like." |
| 6-month follow-up | "I am settled in and plan to continue in the physical activity offer." |
| Reasons for having no experience | |
| 3- and 6-month follow-up | "If no, what is the reason that you haven't tried a physical activity in a civil society organization or similar?" |
| | - "I did not find a physical activity offer that motivated me." |
| | - "I did not find a physical activity offer where I felt safe and welcome." |
| | - "I did not find a physical activity offer where the level fit my needs." |
| | - "I did not find a physical activity offer with a community that fit my needs." |
| | - "I was already active in a physical activity offer in a civil society organization or similar before my municipal program started." |
| | - "I exercise on my own." |
| | - "I am not interested in engaging in physical activities." |

*Digital platform*

| | |
|---|---|
| **Citizens** | |
| Familiarity | |
| Baseline | "On the health center homepage there is an overview of relevant physical activities in the civil society – it looks like this: [screen shot] Have you seen the page before?" |
| **Municipal employees** | |
| Familiarity | |
| Two and six months into the study period | "On the health center homepage there is an overview of relevant physical activities in the civil society – it looks like this: [screen shot] Have you seen the page before?" |
| Use | |
| Two and six months into the study period | "If yes, have you used the homepage in connection with the contact to your citizens?" |
| Relevance | |
| Two and six months into the study period | "If yes, have you found relevant physical activity offers for your citizens on the homepage?" |
| Usefulness | |
| Two and six months into the study period | If yes, is the homepage useful for your work with the citizens?" |
